# Bladder scanning in children: accuracy of volume estimation and detection of urinary retention

**DOI:** 10.64898/2026.11.23.26368740

**Authors:** Anita C. de Jong, Olga E. Arguedas Flores, Job B.M. van Woensel, Kristin Keunen, Geijk V. Eijken, Tanja M. de Kraker, Jolanda M. Maaskant

## Abstract

**Objectives:** Accurate bladder volume assessment supports catheterization decisions in children, but evidence for the accuracy of automated bladder scanners is limited. In a large pediatric cohort, we assessed agreement between bladder scanner-estimated and catheterized volumes, and diagnostic accuracy for detecting urinary retention and elevated post-void residual (PVR).

**Study design:** This prospective cross-sectional study was conducted at a tertiary pediatric hospital. Children aged 0**–**12 years underwent bladder scanning (CUBEScan BioCon-900S) prior to clinically indicated catheterization or cystoscopy. Agreement was assessed using correlation and Bland–Altman analyses. Urinary retention (catheterized volume exceeding expected bladder capacity) and elevated PVR (≥4 years) were defined using age-specific thresholds.

**Results:** In 189 paired measurements from 181 children (median age 2.4 years), correlation was strong (ρ=0.86). Despite a small mean difference (2 mL), 95% limits of agreement were wide (−91 to 95 mL), and only 38% of measurements were within ±20% of catheterized volume. Agreement was moderate in children 2**–**12 years, and lowest in children <2 years and at volumes <100 mL. Overall specificity for urinary retention was 0.88, but sensitivity only 0.75, reflecting underestimation of elevated volumes, with an area under the curve of 0.75 overall versus 0.91 ≥2 years. For elevated PVR, overall sensitivity was 0.98, but specificity 0.63.

**Conclusions:** Bladder scanner-estimated volumes correlated well with catheterized volumes but showed limited agreement, particularly in children <2 years. This device is therefore not recommended in this age group. In older children, it may serve as an adjunct to clinical assessment of urinary retention.

## INTRODUCTION

Accurate assessment of bladder volumes in pediatric patients is essential for diagnosing urinary retention and evaluating post-void residual (PVR).^1, 2^ Catheterization is the gold standard but is invasive and associated with patient discomfort and an increased risk of urinary tract infection.^3^ Conventional bladder ultrasonography provides reliable non-invasive volume estimates, but requires trained staff, and bladder shape-correction formulas are rarely applied in clinical practice.^1, 4-6^ In contrast, portable automated bladder ultrasound devices enable prompt bedside bladder volume assessment, with contemporary devices offering live ultrasound previews and three-dimensional imaging. In adults, published data indicate that most bladder scanners show clinically acceptable agreement with reference methods, with mean differences generally <50 mL and no clear superiority of any single device.^2^ The use of bladder scanners has been shown to reduce catheter-associated urinary tract infections by preventing unnecessary catheterization.^7^

Although bladder scanners are widely used in pediatric clinical practice, evidence regarding their measurement performance in children remains limited. Pediatric studies generally report strong correlations with reference volumes (r = 0.76–0.98).^8-15^ However, mean deviations of 12–33% and wide limits of agreement (LoA) indicate limited individual agreement, using catheterization,^8, 16-19^ infused volumes,^11, 15^ or conventional ultrasound as reference methods.^5, 8, 10, 14, 20^

Agreement is generally poorest in younger children, with age cutoffs ranging from three to seven years across studies, and unreliability in neonates.^10, 15, 17, 18, 21^ However, these age cut-offs lack systematic evaluation. The BioCon-900S incorporates adult and child modes, live preview, and multi-plane imaging, but has been evaluated in only one pediatric study: 29 children aged 0-6 years with spinal dysraphism showed good correlation with infused bladder volume (*r* > 0.9), yet poor agreement in those aged three ≤3 years.^15^

Previous pediatric bladder scanner studies have focused primarily on correlation and agreement metrics, many of which were conducted in highly controlled settings.^5, 8^-^11, 14^-^18, 20, 21^ The clinical applicability of bladder scanners across age groups and their diagnostic accuracy for detecting urinary retention and PVR thresholds have not previously been investigated. Therefore, the primary aim of this study was to evaluate the agreement between BioCon-900S–estimated and catheterized bladder volumes in children aged ≤12 years during routine clinical care. Secondary aims were to evaluate variation in measurement performance by age and bladder volume, and to assess the diagnostic accuracy of the device for detecting urinary retention and PVR thresholds.

## METHODS

### Study design, setting and ethics

This prospective cross-sectional study was conducted in the operating room and pediatric intensive care unit of a tertiary pediatric hospital in the Netherlands between November 1, 2024, and October 15, 2025. Ethical approval was obtained from the institutional review board at the Amsterdam UMC (no 2024.0519). Written informed consent was obtained from parents or legal guardians, an approved opt-out procedure was used for the remaining participants. Data were collected from electronic medical records and case report forms and pseudonymized before analysis. The study was reported in accordance with the STARD guidelines.^22^

### Selection of participants

Children aged zero to twelve years were included if bladder scanning was performed within 15 minutes before clinically indicated urinary catheterization or cystoscopy, yielding complete paired measurements. Exclusion criteria were cystoscopy with irrigation during instrument insertion, structural bladder abnormalities, a history of bladder surgery or hysterectomy, continuous urinary leakage and corrected gestational age less than 37 weeks.

### Index method

Bladder volume estimation using the CUBEScan BioCon-900S (Mcube Technology, Seongnam, South Korea) served as the index method.^23^ Scans were performed by trained healthcare professionals involved in routine patient care, including nurses, pediatric anesthesiologists and pediatric urologists. Patient mode was selected per manufacturer’s instructions: child mode for <25 kg or <120 cm, and adult mode (male or female) for ≥25 kg and ≥120 cm.^23^ When height was unavailable, mode selection was based on weight only. All measurements were included regardless of the mode selected, reflecting routine clinical practice. At least two scans were performed per participant, and the highest displayed bladder volume was used for analysis, consistent with the operator’s manual.^23^

### Reference standard

Bladder volumes obtained by catheterization or cystoscopy served as the reference standard.^17, 18, 21^ Catheterization was performed according to standard clinical protocols, and cystoscopies by pediatric urologists. After urine flow ceased, gentle abdominal compression or syringe aspiration for small volumes was applied to facilitate complete bladder drainage, followed by measurement of the catheterized volume and measurement of any leakage. Blinding between estimated and catheterized volumes was not applied, as catheter drainage served as the objective reference standard.^17, 18, 21^

Urinary retention was defined operationally as a catheterized bladder volume exceeding the expected bladder capacity (EBC), calculated as [*weight in kg* × 7 *ml*] for children <one year and as [(*age in years* + 1) × 30 *ml*] for children ≥one year.^1, 24^ The analyses evaluated whether the bladder scanner accurately identifies volumes exceeding this threshold; clinical signs and symptoms fell outside the scope of a volume-based device.

### Baseline characteristics

Recorded baseline characteristics included age, sex, weight, EBC, reason for admission, presence of neurogenic bladder, procedure location, treatment with intravenous sedation or neuromuscular blocking agents, and indication for catheterization. Descriptive study parameters included bladder scan mode, estimated and catheterized bladder volume, catheter characteristics, operator and time interval between index and reference measurements.

### Statistical analysis

#### Agreement between methods

Association between bladder scanner-estimated and catheterized bladder volumes was visualized using scatterplots and quantified using Pearson’s correlation coefficient (r) or Spearman rank correlation coefficient (ρ), depending on data distribution. Concordance was quantified using the intraclass correlation coefficient (ICC, two-way random-effects model with absolute agreement for single measurements). Mean difference, standard deviations (SD) and 95% LoA (mean ±1.96 × SD) were calculated and expressed as absolute differences (mL). To account for age-related differences in bladder size, volumes were additionally standardized to EBC, with differences expressed as %EBC.^15, 25^ Bland–Altman analysis was used to evaluate agreement across the range of bladder volumes. In the presence of proportional bias, analyses were repeated on log-transformed data after excluding zero volumes. Back-transformed results were reported as geometric mean ratios with 95% LoA. Acceptability was predefined as 95% LoA within ±20% of the catheterized volume. Plots were stratified by urinary retention status.

#### Age- and volume-related performance

Associations of relative differences between methods with catheterized volume and age were assessed using partial Spearman correlations, adjusted for age and catheterized volume, respectively. To evaluate age-dependent measurement performance, Spearman correlations and LoA were calculated across cumulative age groups to identify the age at which strong correlation (ρ >0.90) and acceptable agreement (95% LoA within ±20% of the catheterized volume) were first met. Correlation and agreement metrics were subsequently calculated for children above and below the identified age threshold. To explore volume-dependent agreement, absolute and relative differences were compared with the manufacturer’s phantom-based specifications (<100 mL: ±10 mL; ≥100 mL: ±10%).^23^ In addition, absolute differences were compared across bladder filling categories (%EBC) using boxplots.

#### Diagnostic accuracy for detecting urinary retention and PVR tresholds

Diagnostic accuracy was evaluated for two clinically relevant binary outcomes: urinary retention and age-specific PVR thresholds. Urinary retention was defined as a catheterized volume exceeding the EBC. Age-specific PVR thresholds (>21% of EBC at ≥4–<7 years; >15% of EBC at ≥7–12 years)^1^ were applied to catheterized volumes irrespective of voiding status; the full range of naturally occurring bladder volumes allowed us to evaluate how accurately scanner-estimated volumes distinguish those above from below these thresholds. Scanner-estimated and catheterized volumes obtained as continuous measurements were dichotomized using the same thresholds. Diagnostic performance was quantified using sensitivity, specificity, and the area under the receiver operating characteristic (ROC) curve (AUC) for both outcomes. For urinary retention, cumulative age-specific AUC analyses were additionally performed to identify the youngest age at which excellent discrimination (AUC >0.90) was first achieved.

#### Sample size calculation

The target sample size was 180 children, including at least 100 aged less than three years, based on a Bland– Altman sample size calculation in MedCalc, using a mean difference of 33% EBC, an SD of 22% EBC, a two-sided α of 0.05, and 80% power.^26, 27^

#### General statistical methods

Continuous variables were summarized as mean and SD or median and interquartile range (IQR), and categorical variables as frequencies and percentages. For Spearman ρ and AUCs, 95% CI were calculated using 2,000 resamples. Confidence intervals for proportions were calculated using the Wilson method. Statistical analyses were performed using R version 4.4.3.^28^

## RESULTS

### Participants and baseline characteristics

Of 224 patients screened for eligibility, 181 were included, contributing to 189 bladder volume measurements (Fig 1). Patient, procedural, and measurement characteristics are presented in Table 1.

**Table 1.**
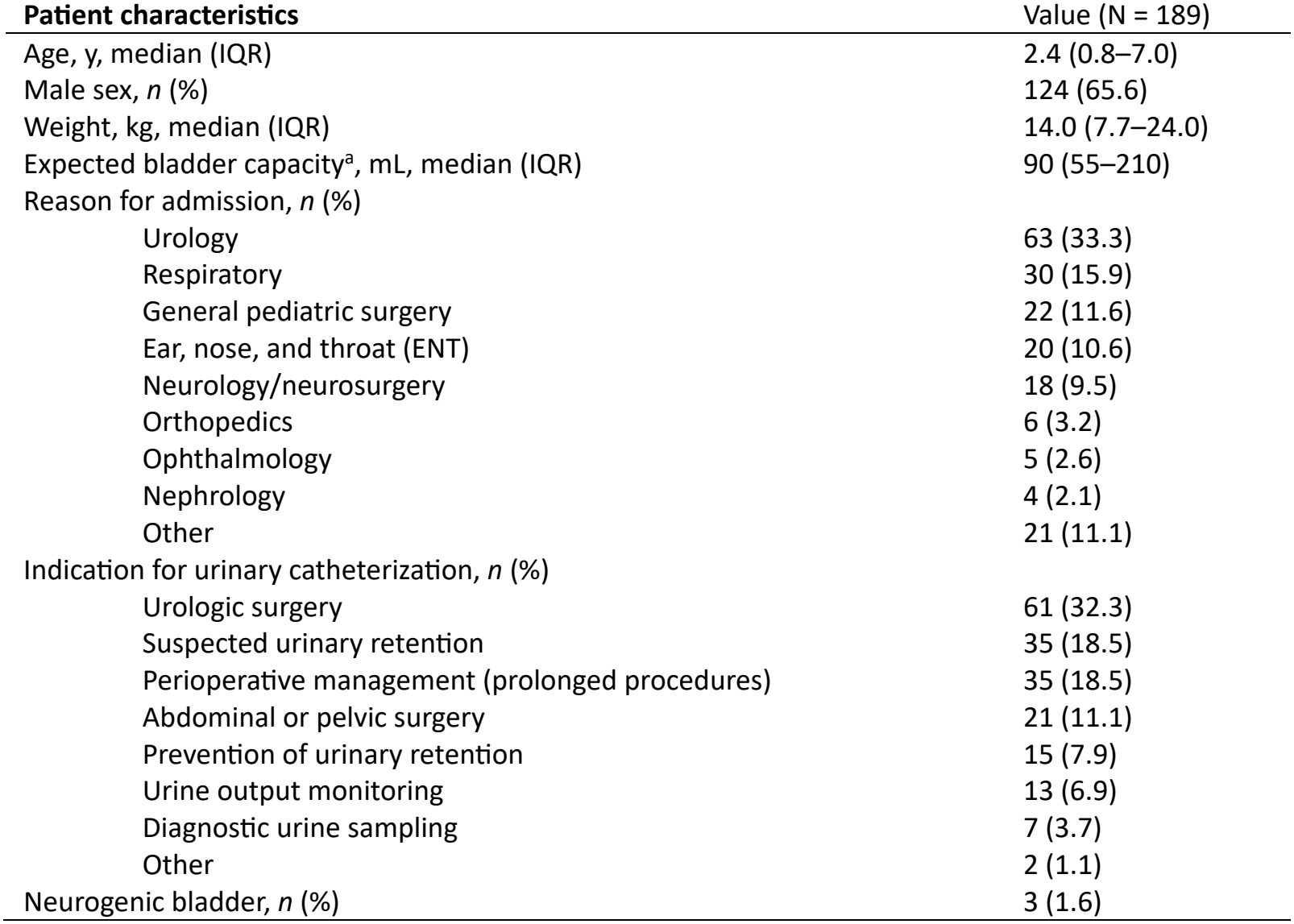
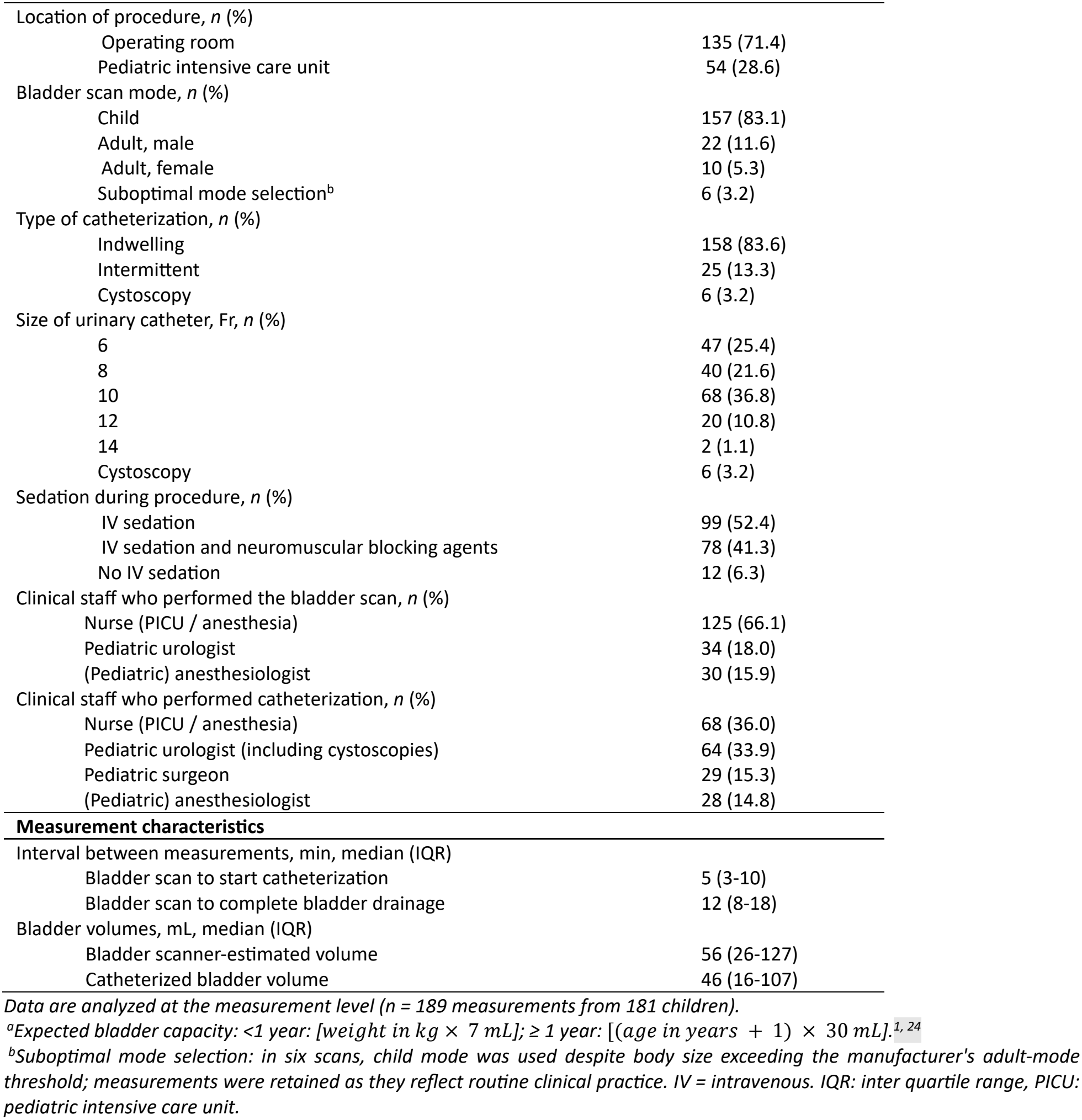
Patient, procedural, and measurement characteristics.

| **Patient characteristics** | Value (N = 189) |
|---|---|
| Age, y, median (IQR) | 2.4 (0.8–7.0) |
| Male sex, *n* (%) | 124 (65.6) |
| Weight, kg, median (IQR) | 14.0 (7.7–24.0) |
| Expected bladder capacity<sup>a</sup>, mL, median (IQR) | 90 (55–210) |
| Reason for admission, *n* (%) |  |
| Urology | 63 (33.3) |
| Respiratory | 30 (15.9) |
| General pediatric surgery | 22 (11.6) |
| Ear, nose, and throat (ENT) | 20 (10.6) |
| Neurology/neurosurgery | 18 (9.5) |
| Orthopedics | 6 (3.2) |
| Ophthalmology | 5 (2.6) |
| Nephrology | 4 (2.1) |
| Other | 21 (11.1) |
| Indication for urinary catheterization, *n* (%) |  |
| Urologic surgery | 61 (32.3) |
| Suspected urinary retention | 35 (18.5) |
| Perioperative management (prolonged procedures) | 35 (18.5) |
| Abdominal or pelvic surgery | 21 (11.1) |
| Prevention of urinary retention | 15 (7.9) |
| Urine output monitoring | 13 (6.9) |
| Diagnostic urine sampling | 7 (3.7) |
| Other | 2 (1.1) |
| Neurogenic bladder, *n* (%) | 3 (1.6) |
| Location of procedure, *n* (%) |  |
| Operating room | 135 (71.4) |
| Pediatric intensive care unit | 54 (28.6) |
| Bladder scan mode, *n* (%) |  |
| Child | 157 (83.1) |
| Adult, male | 22 (11.6) |
| Adult, female | 10 (5.3) |
| Suboptimal mode selection<sup>b</sup> | 6 (3.2) |
| Type of catheterization, *n* (%) |  |
| Indwelling | 158 (83.6) |
| Intermittent | 25 (13.3) |
| Cystoscopy | 6 (3.2) |
| Size of urinary catheter, Fr, *n* (%) |  |
| 6 | 47 (25.4) |
| 8 | 40 (21.6) |
| 10 | 68 (36.8) |
| 12 | 20 (10.8) |
| 14 | 2 (1.1) |
| Cystoscopy | 6 (3.2) |
| Sedation during procedure, *n* (%) |  |
| IV sedation | 99 (52.4) |
| IV sedation and neuromuscular blocking agents | 78 (41.3) |
| No IV sedation | 12 (6.3) |
| Clinical staff who performed the bladder scan, *n* (%) |  |
| Nurse (PICU / anesthesia) | 125 (66.1) |
| Pediatric urologist | 34 (18.0) |
| (Pediatric) anesthesiologist | 30 (15.9) |
| Clinical staff who performed catheterization, *n* (%) |  |
| Nurse (PICU / anesthesia) | 68 (36.0) |
| Pediatric urologist (including cystoscopies) | 64 (33.9) |
| Pediatric surgeon | 29 (15.3) |
| (Pediatric) anesthesiologist | 28 (14.8) |
| **Measurement characteristics** |  |
| Interval between measurements, min, median (IQR) |  |
| Bladder scan to start catheterization | 5 (3-10) |
| Bladder scan to complete bladder drainage | 12 (8-18) |
| Bladder volumes, mL, median (IQR) |  |
| Bladder scanner-estimated volume | 56 (26-127) |
| Catheterized bladder volume | 46 (16-107) |

*Data are analyzed at the measurement level (n = 189 measurements from 181 children).*
<sup>a</sup>*Expected bladder capacity: <1 year: [weight in kg × 7 mL]; ≥ 1 year: [(age in years + 1) × 30 mL].*<sup>*1, 24*</sup>
<sup>b</sup>*Suboptimal mode selection: in six scans, child mode was used despite body size exceeding the manufacturer's adult-mode threshold; measurements were retained as they reflect routine clinical practice. IV = intravenous. IQR: inter quartile range, PICU: pediatric intensive care unit.*

**Fig 1.**
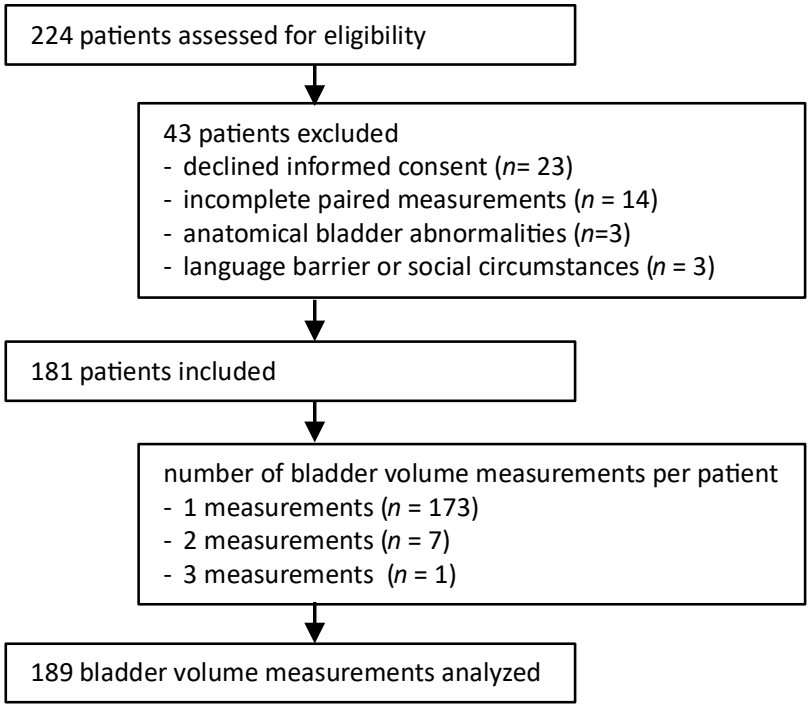
Flowchart of participant inclusion and measurement selection.

### Agreement between methods

A strong positive association was observed between bladder scanner-estimated and catheterized volumes, with greater variability and a tendency toward underestimation at higher bladder volumes, particularly in children with urinary retention (Fig 2). Correlation was strong and reliability was good (Table 2). The mean absolute difference was small (2.1 mL). However, the LoA were wide (−90.7 to 95.0 mL), indicating limited precision at the individual level, particularly in children <2 years old. Bland–Altman analysis on the absolute scale showed increasing variability with increasing bladder volume, with similar findings after standardization to %EBC (Figs S1 and S2). Log-transformed Bland–Altman analysis demonstrated proportional bias, with overestimation of catheterized volumes <100 mL and underestimation >200 mL, most pronounced in children ≥2 years old with urinary retention (Fig. 3). Only 69 of 180 measurements (38.3%; 95% CI, 31.5%–45.6%) fell within the predefined ±20% acceptability margin compared to catheterized volume, after exclusion of zero values (n=9). This proportion was lower in children <2 years (24/83, 28.9%) than in those ≥2 years (45/97, 46.4%).

**Table 2.** Bladder scanner-estimated versus catheterized bladder volumes: correlation, reliability, and agreement.

| **Age** | ***N*** | **Spearman ρ**<br>**(95% CI)** | **ICC**<br>**(95% CI)** | **Mean difference, mL**<br>**(SD, 95% LoA)** | **Mean difference, %EBC**<br>**(SD, 95% LoA),** | **Geometric ratio**<br>**(95% LoA)** |
|---|---|---|---|---|---|---|
| 0–12 years | 189 | 0.86 (0.80–0.91) | 0.88 (0.84–0.91) | 2.14 (47.4, −90.7 to 95.0) | 11.84 (56.6, −99 to 122.7) | 1.26 (0.3 – 5.35) |
| 0–2 years | 88 | 0.65 (0.46–0.80) | 0.79 (0.69–0.86) | 5.75 (28.2; −49.6 to 61.1) | 23.93 (75.8, −124 to 172.5) | 1.34 (0.21 – 8.66) |
| ≥2–12 years | 101 | 0.93 (0.88–0.96) | 0.86 (0.80–0.90) | −1.01 (59.2, −117.1 to 115.0) | 1.30 (7.9, −53.4 to 56) | 1.19 (0.46 – 3.09) |

*CI: confidence interval. %EBC: bladder volumes standardized to percentage of expected bladder capacity. ICC: intraclass correlation coefficient based on a two-way random-effects model with absolute agreement for single measurements [2,1]. Geometric ratio represents the estimated-to-catheterized bladder volume ratio based on log-transformed Bland–Altman analysis.*

**Fig 2.**
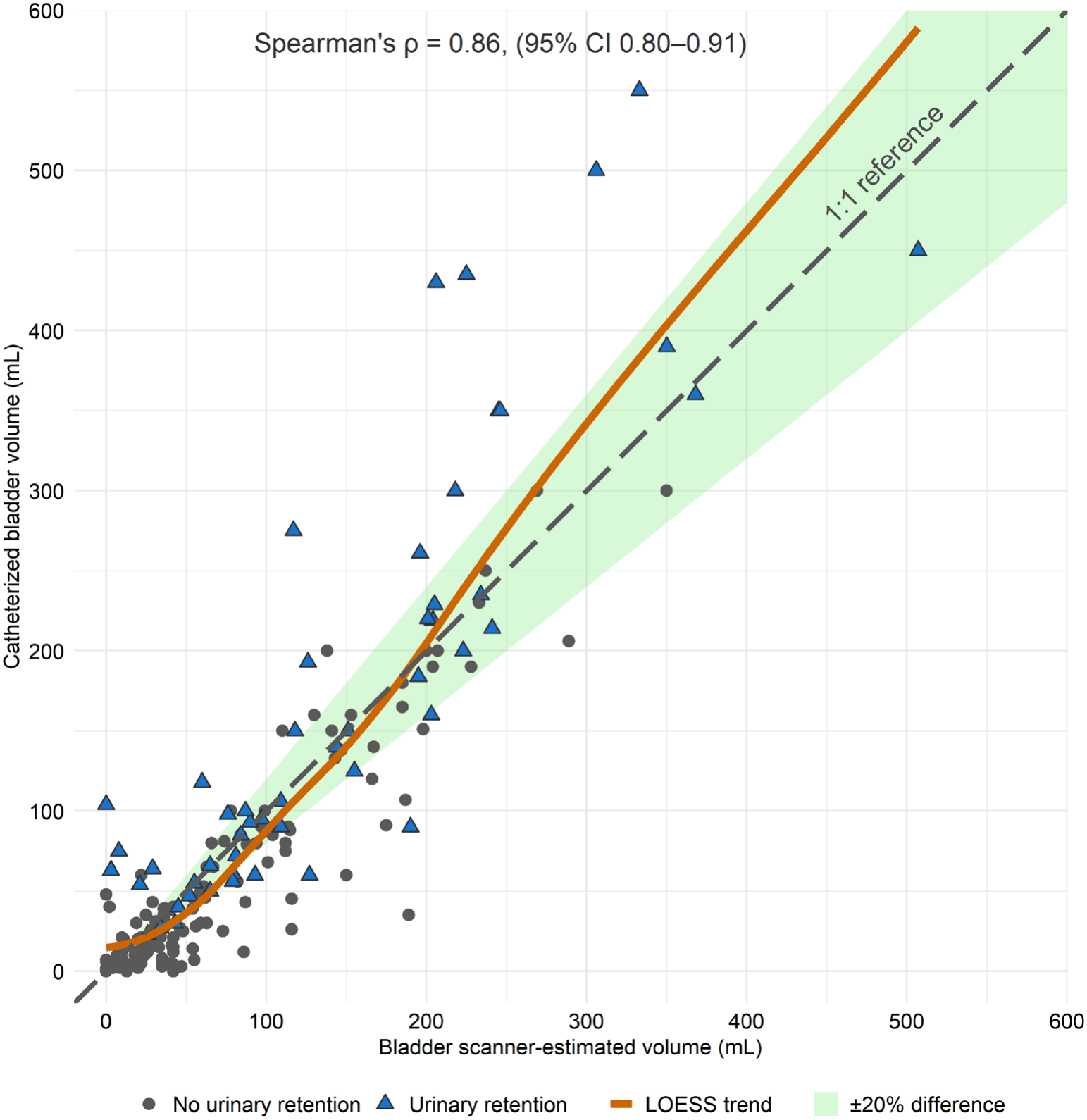
Scatterplot of bladder scanner-estimated versus catheterized bladder volumes. Symbols distinguish measurements without and with urinary retention; lines indicate the 1:1 reference, LOESS trend, and ±20% difference.

**Fig 3.**
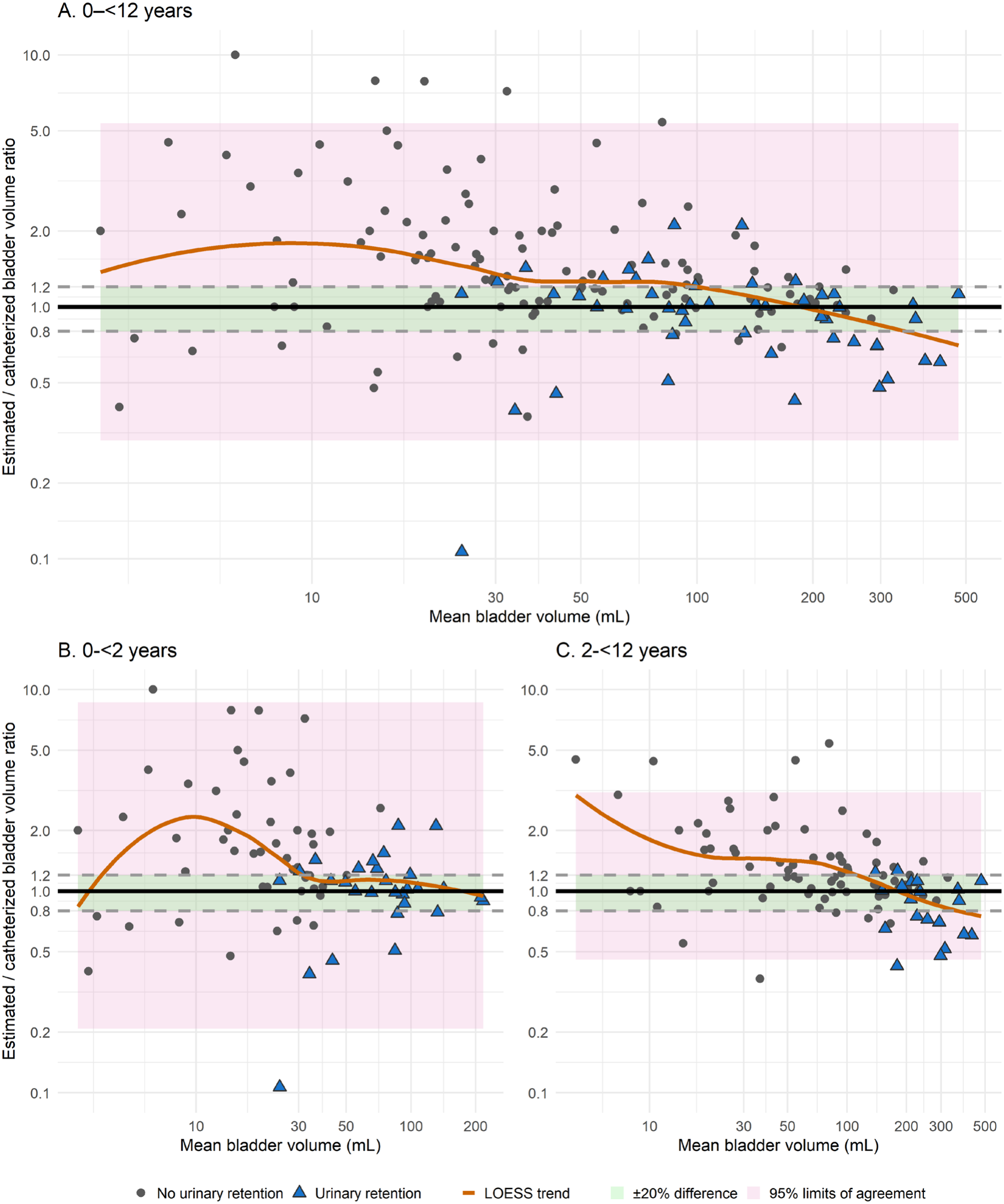
Bland–Altman plots of bladder scanner-estimated versus catheterized bladder volume, overall and by age group. A, children 0–<12 years; (*n* = 189); B, children <2 years (*n* = 88); C, children ≥2– <12 years (*n* = 101). *The x-axis shows geometric mean bladder volume (mL, logarithmic scale), and the y-axis the estimated-to-catheterized volume ratio (logarithmic scale)*.

### Age- and volume-related performance

Smaller bladder volumes remained independently associated with larger relative differences between scanner-estimated and catheterized volumes (partial ρ = −0.49, *P* <0.001), consistent with the greater variability observed at volumes <100 mL (Fig 3A). Age was not independently associated with relative differences (partial ρ = 0.05, *P* = 0.50). Correlations nevertheless improved across age groups, remaining above ρ = 0.9 ≥2 years (Table S1). The LoA narrowed from approximately two years onward, but no clear age threshold was identified at which the predefined ±20% agreement criterion was consistently met (Fig 3C and Fig S3).

Compared with manufacturer-specified accuracy, the mean absolute difference for volumes <100 mL slightly exceeded the ±10 mL threshold, whereas the mean relative difference for volumes ≥100 mL remained within ±10% threshold (Table 3). However, the LoA markedly exceeded these specifications in both absolute and relative volumes. Across bladder filling categories (%EBC), median absolute differences were small. The device showed slight overestimation at levels <100% EBC (Fig. 4). At filling levels ≥100% EBC (urinary retention), variability increased substantially, with underestimations to a greater extent than overestimations.

**Table 3.** Agreement between bladder scanner-estimated and catheterized bladder volumes: comparison with manufacturer accuracy specifications.

| **Catheterized bladder volume** | **Mean difference, mL, mean (SD, 95% LoA)** | **Mean relative difference, %, mean (SD, 95% LoA)** | **Manufacturer criterion<sup>a</sup>** |
|---|---|---|---|
| <100 mL | 12.5 (27.7, −41.7 to 66.8) | 73.5 (155.7, −231.7 to 378.7) | ± 10 mL |
| ≥100 mL | −24.5 (71.5, −164.8 to 115.7) | −6.5 (28.6, −62.5 to 49.5) | ± 10 % |

<sup>a</sup> *Manufacturer-specified phantom-based deviation criteria are ±10 mL for volumes <100 mL and ±10% for volumes ≥100 mL.<sup>23</sup>*

**Fig 4.**
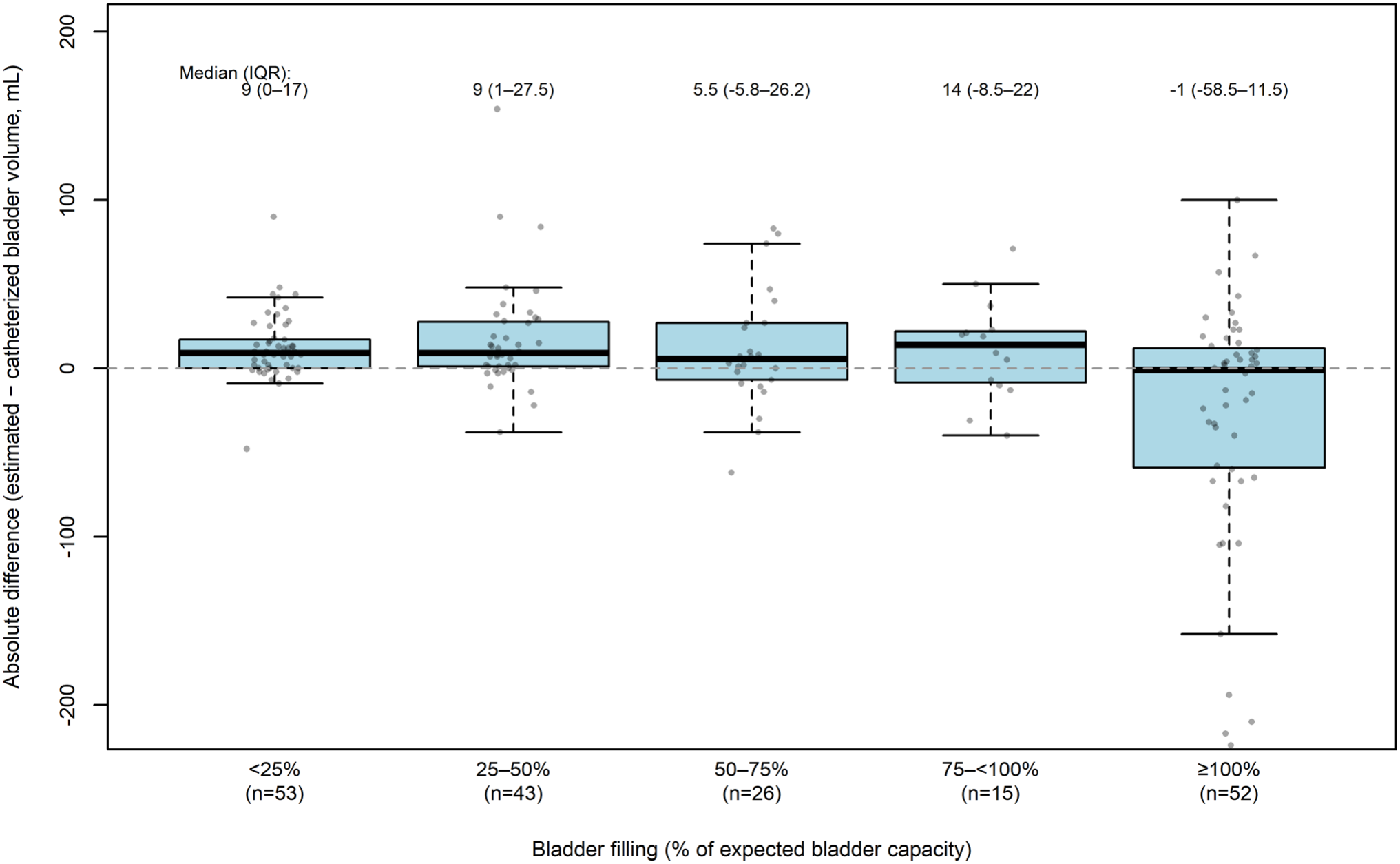
Absolute difference between bladder scanner-estimated and catheterized bladder volumes by bladder filling category. Bladder filling was defined according to catheterized bladder volume as percentage of expected bladder capacity (EBC, estimated as: <1 year: [weight in kg × 7 ml]; ≥ 1 year: [(age in years + 1) × 30 ml].^1, 24^. Boxes represent the median and interquartile range, whiskers extend to 1.5 × IQR; values beyond are plotted as outliers.

### Diagnostic accuracy for detecting urinary retention

Catheterized volumes exceeded the EBC in 52 of 189 catheterizations (27.5%), of which 35 were performed for suspected urinary retention and 17 for other indications. Diagnostic accuracy for detecting urinary retention at the predefined threshold varied by age group. In children aged ≥2 years, discrimination was excellent, with high specificity but moderate sensitivity (Table 4). In children <2 years, sensitivity was higher, but specificity was lower. The contingency table, AUC-analyses across cumulative age groups, and ROC curves are shown in Tables S4, S5 and Fig S5.

**Table 4.** Diagnostic accuracy of bladder scanner–estimated volumes for detecting urinary retention.

| Age group | *n* | Urinary retention<sup>a</sup> *n*(%) | Sensitivity (95% CI) | Specificity (95% CI) | AUC (95% CI) |
|---|---|---|---|---|---|
| 0–<2 years | 88 | 31 (35.2%) | 0.84 (0.67–0.93) | 0.75 (0.63–0.85) | 0.82 (0.71–0.93) |
| ≥2–12 years | 101 | 21 (20.8%) | 0.62 (0.41–0.79) | 0.96 (0.90–0.99) | 0.91 (0.85–0.96) |
| 0–12 years | 189 | 52 (27.5%) | 0.75 (0.61–0.86) | 0.88 (0.81–0.92) | 0.75 (0.67–0.83) |

*<sup>a</sup> Urinary retention: catheterized bladder volume ≥ expected bladder capacity, defined as [weight in kg× 7 mL] for children aged <one year and as [(age in years + 1) × 30 mL] for children ≥one year.<sup>1, 24</sup>*

### Diagnostic accuracy at age-specific PVR thresholds

In children aged four years and older, catheterized volumes exceeded PVR thresholds in 62/81 measurements (76.5%). Sensitivity of bladder scanner–estimated volumes for detecting elevated PVR thresholds was high in both age groups (≥4–<7 years: 1.00, 95% CI 0.88–1.00; ≥7–<12 years: 0.97, 95% CI 0.85–0.99), whereas specificity was low (0.57, 95% CI 0.25–0.84 and 0.67, 95% CI 0.39–0.86, respectively; Table S4). Corresponding AUCs indicated excellent discrimination in both age groups: ≥4– <7 years 0.98 (95% CI 0.95–1.00), ≥7–<12 years 0.94 (95% CI 0.87–1.00). The contingency table and ROC curves are shown in Table S5 and Fig S5.

## DISCUSSION

Bladder scanner-estimated volumes correlated strongly with catheterized bladder volumes in children <12 years, with a small absolute mean difference. However, individual agreement was limited, reflected by wide LoA and inconsistent conformity to the predefined ±20% criterion. Agreement was lowest in children <2 years and at bladder volumes <100 mL, with overestimation at smaller bladder volumes and underestimation at larger volumes. Overestimation at low volumes reduced specificity for detecting elevated PVR, whereas underestimation at high volumes reduced sensitivity for detecting urinary retention.

Despite good overall discrimination in children aged ≥2 years, sensitivity at the clinical threshold was only moderate.

This pattern of proportional bias is consistent with a review of adult studies.^2^ Reported mean differences of <50 mL are considered acceptable in adults, but in children such deviations constitute a substantially larger proportion of bladder capacity. Several devices have been evaluated in children,^5, 9^-^12, 14, 15, 17-21^ however, this device has previously been assessed in only one pediatric cohort.^13^ In that study, performance was considered acceptable in children >3 years, based on a mean bias of <30% EBC. In contrast, we assessed clinical acceptability against a predefined criterion of agreement within ±20% of the catheterized volume. This criterion was not met, indicating that individual volume estimates lack clinically acceptable accuracy. For this device, the mean difference (%EBC) was smaller than previously reported and generally within the manufacturer’s specifications.^15, 23^ This indicates that, on average, scanner estimates were close to catheterized volumes. However, LoA were wider than both the manufacturer’s specifications and previously reported values, likely reflecting the greater variability of routine clinical practice compared to highly controlled settings.^15, 23^ Comparable findings have been reported for other portable scanners in children: small mean differences with wide LoA in some studies,^10, 17, 18^ and conclusions of reliability without agreement analysis in others.^14^

Our systematic age-stratified analysis identified reduced performance primarily <2 years of age, consistent with previous studies reporting reduced performance in young children.^10, 14, 15, 17, 18, 21^ Although these studies proposed higher age thresholds of three to seven years, none systematically evaluated the influence of age on scanner performance, and the rationale for their chosen thresholds was not reported. Likewise, we and other studies found reduced accuracy at bladder volumes <50–100 mL.^11, 17, 18, 20, 29^ Because younger children generally have smaller bladder volumes, the effects of age and bladder volume are difficult to disentangle. However, our adjusted analyses showed that smaller bladder volume, rather than age, was independently associated with lower agreement. This may explain the low specificity, but high sensitivity observed for detecting elevated PVR, as small measurement differences have a greater impact at low bladder volumes. In addition, anatomical and technical factors, such as a more intra-abdominal bladder position and the relatively large probe size, may further limit the accuracy of bladder scanner measurements in young children.^25^

At higher bladder volumes, the opposite pattern was observed. In children ≥2 years, sensitivity for detecting urinary retention was only moderate, whereas both specificity and AUC were high. A high AUC indicates good overall discrimination but does not guarantee adequate sensitivity at the clinical threshold, as it reflects performance across all possible thresholds. In our study, scanner-estimated volumes below the clinical threshold did not reliably exclude urinary retention, suggesting that bladder scanning may be more useful for confirming than for excluding urinary retention. This is consistent with a previous study reporting that not all children with bladder volumes exceeding the EBC were identified.^13^

Several limitations of this study should be considered. First, child mode was selected for six scans despite the manufacturer recommending adult mode above specified body size thresholds. These measurements were deliberately retained, as they reflect routine clinical practice. Second, in young children, small-diameter catheters may not always have completely emptied the bladder, possibly leaving a small residual volume despite syringe aspiration and gentle abdominal compression, underestimating the true bladder volume. Although small in absolute terms, such deviations may disproportionately affect measurements at low bladder volumes by both catheterization and bladder scanning, and may partly explain the limited performance in young children. Third, ongoing urine production during catheterization may have inflated the catheterized volumes. This effect is likely limited: in small bladders, catheterization is completed before substantial urine production occurs, whereas in large bladders, additional urine production is likely small relative to bladder volume. Fourth, most measurements were obtained in the operating room or pediatric intensive care unit, predominantly in sedated or anesthetized children; device performance in awake children may differ and requires further study. Finally, consistent with previous studies, we evaluated scanner-estimated volumes only, not the underlying scan acquisition or image interpretation.

Despite these limitations this is, our cohort is among the largest in pediatric bladder scanner research, enabling systematic evaluation of age- and bladder volume-specific performance. Beyond confirming and refining previously reported limitations in children < 2 years and at bladder volumes <100 mL,^11, 17, 18, 20, 29^ we determined the diagnostic accuracy of the device for detecting urinary retention and elevated PVR. To our knowledge, these clinically relevant outcomes have not previously been assessed in pediatric bladder scanner studies. This was possible because all catheterized children were included irrespective of indication, capturing the full range of naturally occurring bladder volumes, including elevated volumes in children without suspected urinary retention. As the device was evaluated by multiple operators in routine clinical practice, our findings are likely generalizable to comparable settings.

Our findings have important implications for clinical use of this bladder scanner in children. Given the limited agreement in children under two years of age, this device should is not recommended in this age group. In older children, scanner-estimated volumes should be interpreted with caution, particularly at smaller volumes. In children aged ≥2 years, this device may assist in the evaluation of urinary retention, although larger volumes may be underestimated. Therefore, displayed volumes below the defined threshold do not reliably exclude urinary retention. Estimated volumes should therefore always be interpreted in the context of clinical signs and symptoms, such as a palpable bladder, lower abdominal pain or discomfort, and voiding difficulties. As such, this bladder scanner may support decision-making regarding urinary catheterization as an adjunct to, rather than a substitute for, clinical evaluation.

Further research should establish device performance in awake children and investigate the impact of scan acquisition, image interpretation, and scan mode selection. Such research should inform improvements in pediatric bladder scanner technology and clinical guidance on its appropriate use.

## Data Availability

All data produced in the present study are available upon reasonable request to the authors

## Abbreviations

AUC: area under the curve
EBC: expected bladder capacity
IQR: interquartile range
LoA: limits of agreement
PVR: post-void residual
ROC: receiver operating characteristic
SD: standard deviation

## Author contributions

**Anita C. de Jong:** Conceptualization, Methodology, Project Administration, Resources, Software, Investigation, Data Curation, Formal Analysis, Validation, Visualization, Writing – original draft, Writing – review & editing. **Olga E. Arguedas Flores:** Conceptualization, Methodology, Investigation, Validation, Writing – review & editing. **Job B.M. van Woensel:** Conceptualization, Methodology, Validation, Supervision, Writing – review & editing. **Kristin Keunen:** Conceptualization, Investigation, Writing – review & editing. **Geijk V. Eijken:** Investigation, Project Administration, Writing – review & editing. **Tanja M. de Kraker:** Conceptualization, Writing – review & editing. **Jolanda M. Maaskant:** Conceptualization, Methodology, Validation, Supervision, Writing – review & editing. All authors approved the final manuscript as submitted and agree to be accountable for all aspects of the work.

## Acknowledgements

We sincerely thank the children and their parents or caregivers for participation in this study. We also thank the healthcare professionals who contributed to data collection.

## Data availability statement

The data that support the findings of this study are available on request from the corresponding author. The data are not publicly available due to privacy or ethical restrictions.

## Declaration of generative AI and AI-assisted technologies in the manuscript preparation process

During the preparation of this work, the first author (AdJ) used the institutional, proprietary Tulp Chat Amsterdam UMC (GPT-5.3) for language editing and support for statistical coding. The author reviewed, verified, and edited all output and takes full responsibility for the content of the published article.^30^

## Notes

***Conflicts of interest statement:*** The authors have no conflicts of interest relevant to this article to disclose.

**Funding / support**: This research did not receive any specific grant from funding agencies in the public, commercial, or not-for-profit sectors. The bladder scanners used in this study were part of the institution’s routine clinical equipment. No equipment or other support was provided by the manufacturer.

### Competing Interest Statement

The authors have declared no competing interest.

### Author Declarations

Ethical approval was obtained from the institutional review board at the Amsterdam UMC (no 2024.0519).

