## Supplemental Materials for "Bladder scanning in children: accuracy of volume estimation and detection of urinary retention"

***PREPRINT – not peer reviewed***

*Anita C. de Jong, MSc[a,b]; Olga E. Arguedas Flores, MD[c]; Job B.M. van Woensel, MD, PhD[a,b]; Kristin Keunen, MD, PhD[d]; Geijk V. Eijken, MSc[e]; Tanja M. de Kraker, MSc[c]; Jolanda M. Maaskant, PhD[b,f]*

### Affiliations

[a] Department of Pediatric Intensive Care, Emma Children's Hospital, Amsterdam UMC Location University of Amsterdam, Amsterdam, The Netherlands

[b] Amsterdam Reproduction and Development research institute, Amsterdam, The Netherlands

[c] Department of Pediatric Urology, Emma Children's Hospital, Amsterdam UMC Location University of Amsterdam, Amsterdam, The Netherlands

[d] Department of Anesthesiology, Amsterdam UMC Location University of Amsterdam, Amsterdam, The Netherlands

[e] Department of Radiology and Nuclear Medicine, Amsterdam UMC Location University of Amsterdam, Amsterdam, The Netherlands

[f] Emma Children's Hospital, Amsterdam UMC Location University of Amsterdam, Amsterdam, The Netherlands

### SUPPLEMENTAL TABLES

- Supplemental Table 1. Spearman correlation between bladder scanner-estimated and catheterized bladder volumes across cumulative age groups
- Supplemental Table 2. 2×2 Contingency table for bladder scanner-based classification of urinary retention compared with catheterized bladder volume
- Supplemental Table 3. AUC- analyses for bladder scanner-based discrimination of urinary retention across cumulative age groups
- Supplemental Table 4. Diagnostic accuracy of bladder scanner based classifications at PVR thresholds in children aged ≥4 years, compared with catheterized bladder volume
- Supplemental Table 5. 2×2 Contingency table for accuracy of bladder scanner-based classification at PVR thresholds compared with catheterized bladder volume

### SUPPLEMENTAL FIGURES

- Supplemental Fig 1. Bland–Altman plot of absolute differences between bladder scanner-estimated and catheterized bladder volumes
- Supplemental Fig 2. Bland–Altman plot of the relative differences between bladder scanner-estimated and catheterized bladder volumes, expressed as percentage of expected bladder capacity
- Supplemental Fig 3. Limits of agreement for bladder scanner-estimated versus catheterized bladder volumes across cumulative age groups
- Supplemental. Fig 4. ROC curves for bladder scanner-based detection of urinary retention
- Supplemental. Fig 5. ROC curves for bladder scanner-based detection of elevated post-void residual

### Supplemental Table 1. Spearman correlation between bladder scanner-estimated and catheterized bladder volumes across cumulative age groups

| Age threshold (years) | *N* | Spearman ρ (95% CI) |
|---|---|---|
| ≥0 | 189 | 0.86 (0.79–0.91) |
| ≥1 | 134 | 0.89 (0.83–0.94) |
| ≥2 | 101 | 0.93 (0.88–0.96) |
| ≥3 | 89 | 0.92 (0.86–0.95) |
| ≥4 | 81 | 0.93 (0.88–0.95) |
| ≥5 | 74 | 0.93 (0.88–0.95) |
| ≥6 | 60 | 0.93 (0.88–0.96) |
| ≥7 | 47 | 0.93 (0.87–0.96) |
| ≥8 | 40 | 0.92 (0.84–0.95) |
| ≥9 | 31 | 0.91 (0.81–0.96) |
| ≥10 | 19 | 0.86 (0.58–0.96) |
| ≥11 | 7 | Not calculated (n < 10) |

*Spearman ρ = Spearman correlation coefficient. Confidence intervals were estimated using bootstrap resampling (2000 iterations). Correlations were calculated cumulatively for children aged equal to or greater than each age threshold. Correlations and 95% confidence intervals were not calculated when n <10.*

**Supplemental Table 2. 2×2 Contingency table for bladder scanner-based classification of urinary retention compared with catheterized bladder volume**

| Bladder scanner-estimated volume | Catheterized bladder volume ≥100% EBC | Catheterized bladder volume <100% EBC |
|---|---|---|
| ≥100% EBC | TP = 39 | FP = 17 |
| <100% EBC | FN = 13 | TN = 120 |

*TP: true positive; FP: false positive; FN: false negative; TN: true negative. Urinary retention was defined as a catheterized bladder volume ≥100% expected bladder capacity (EBC): <1 year:* $[weight\ in\ kg \times 7\ mL]$*; ≥ 1 year:* $[(age\ in\ years + 1) \times 30\ mL]$*.*[1,2]

### Supplemental Table 3. AUC-analyses for bladder scanner-based discrimination of urinary retention across cumulative age groups

| Age threshold (years) | *N* | Urinary retention, *n* | AUC (95% CI) |
|---|---|---|---|
| ≥0 years | 189 | 52 | 0.75 (0.67–0.83) |
| ≥1 years | 134 | 31 | 0.80 (0.70–0.90) |
| ≥2 years | 101 | 21 | 0.91 (0.85–0.96) |
| ≥3 years | 89 | 20 | 0.89 (0.82–0.96) |
| ≥4 years | 81 | 19 | 0.89 (0.82–0.96) |
| ≥5 years | 74 | 16 | 0.91 (0.84–0.98) |
| ≥6 years | 60 | 10 | 0.93 (0.86–0.99) |
| ≥7 years | 47 | 7 | 0.93 (0.86–1.00) |
| ≥8 years | 40 | 5 | 0.93 (0.84–1.00) |
| ≥9 years | 31 | 5 | 0.90 (0.78–1.00) |
| ≥10 years | 19 | 5 | 0.83 (0.64–1.00) |
| ≥11 years | 7 | 1 | Not calculated (n < 10) |

*AUC = Area Under receiver operator Curve. Urinary retention was defined as a catheterized bladder volume ≥100% expected bladder capacity (EBC), defined as [weight in kg× 7 mL] for children aged <one year and as [(age in years + 1) × 30 mL] for children ≥one year.*[1,2] *AUC was not calculated when n<10 measurements.*

### Supplemental Table 4. Diagnostic accuracy of bladder scanner based classifications at PVR thresholds in children aged ≥4 years, compared with catheterized bladder volume

| Age group | *n* | Elevated PVR, *n*(%) | Sensitivity (95% CI) | Specificity (95% CI) | AUC (95% CI) |
|---|---|---|---|---|---|
| 4–<7 years | 34 | 27 (79.4%) | 1.00 (0.88–1.00) | 0.57 (0.25–0.84) | 0.98 (0.95–1.00) |
| 7–12 years | 47 | 35 (74.5%) | 0.97 (0.85–0.99) | 0.67 (0.39–0.86) | 0.94 (0.87–1.00) |
| 4–12 years | 81 | 62 (76.5%) | 0.98 (0.91–1.00) | 0.63 (0.41–0.81) | 0.95 (0.90–1.00) |

*AUC: area under the receiver operating characteristic curve. PVR: bladder volumes exceeding post-void residual thresholds, defined as >21% of expected bladder capacity (EBC) for children aged ≥4–<7 years, and >15% for children aged ≥7–<12 years, irrespective of voiding status.*[2]

**Supplemental Table 5. 2×2 Contingency table for accuracy of bladder scanner-based classification at PVR thresholds compared with catheterized bladder volume**

| | Catheterized volume exceeding PVR threshold | Catheterized volume exceeding PVR threshold − |
|---|---|---|
| Bladder scanner-estimated volume exceeding PVR threshold + | TP = 61 | FP = 7 |
| Bladder scanner-estimated volume exceeding PVR threshold − | FN = 1 | TN = 12 |

*TP: true positive; FP: false positive; FN: false negative; TN: true negative. PVR: bladder volumes exceeding post-void residual thresholds, defined as >21% of expected bladder capacity (EBC) for children aged ≥4–<7 years, and >15% for children aged ≥7–<12 years, irrespective of voiding status.*[2]

**Supplemental Fig 1. Bland–Altman plot of absolute differences between bladder scanner-estimated and catheterized bladder volumes**

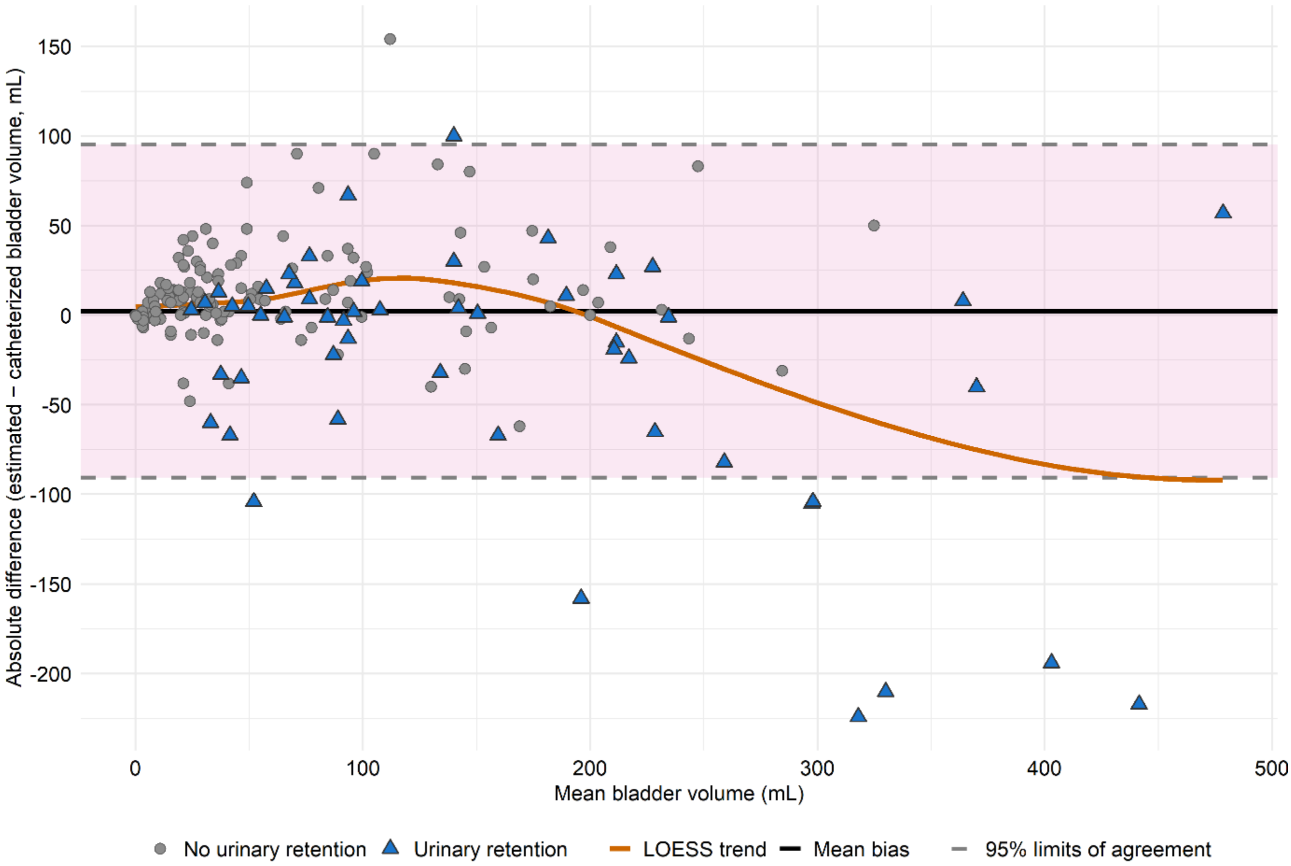

**Supplemental Fig 2. Bland–Altman plot of the relative differences between bladder scanner-estimated and catheterized bladder volumes, expressed as percentage of expected bladder capacity**

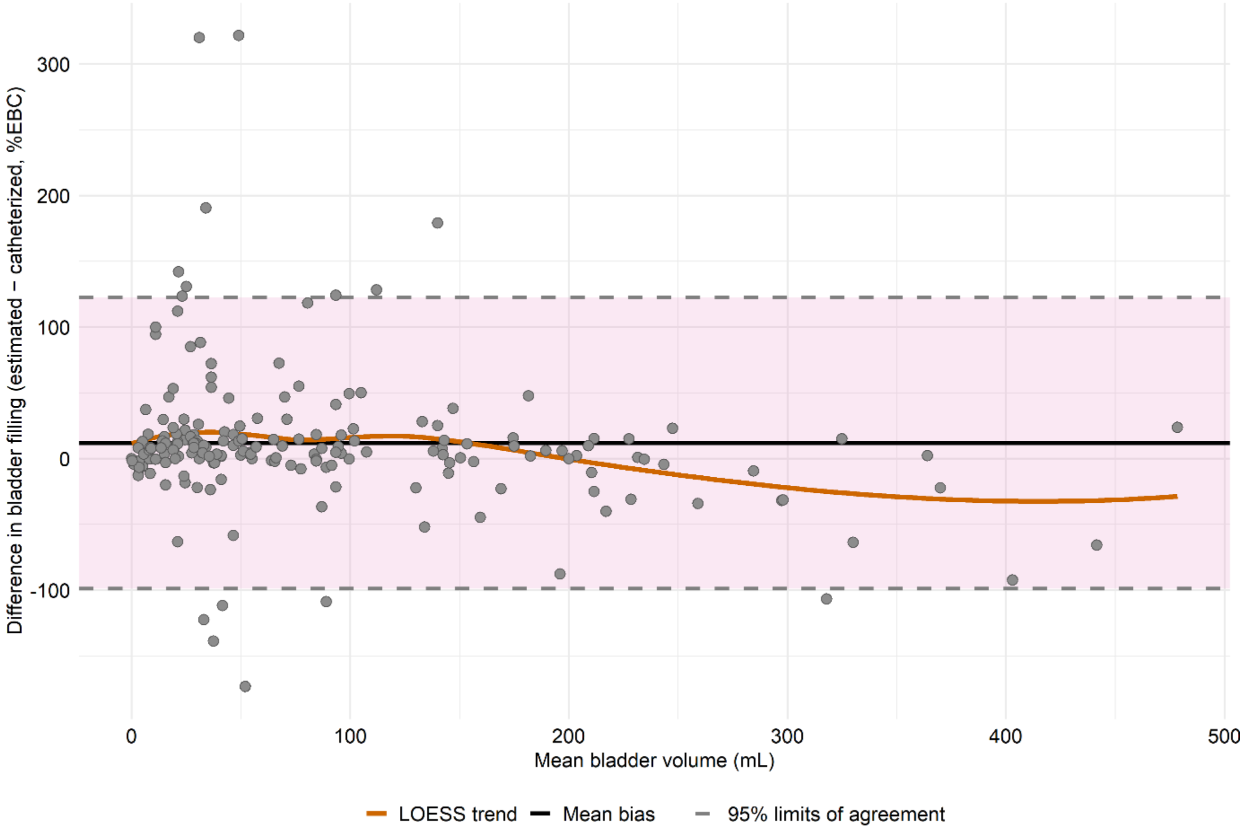


*Differences represent estimated minus catheterized bladder filling expressed as percentage of expected bladder capacity (%EBC). The black line indicates mean bias; dashed lines indicate 95% limits of agreement; the orange line represents LOESS smoothing.*

**Supplemental Fig 3. Limits of agreement for bladder scanner-estimated versus catheterized bladder volumes across cumulative age groups**

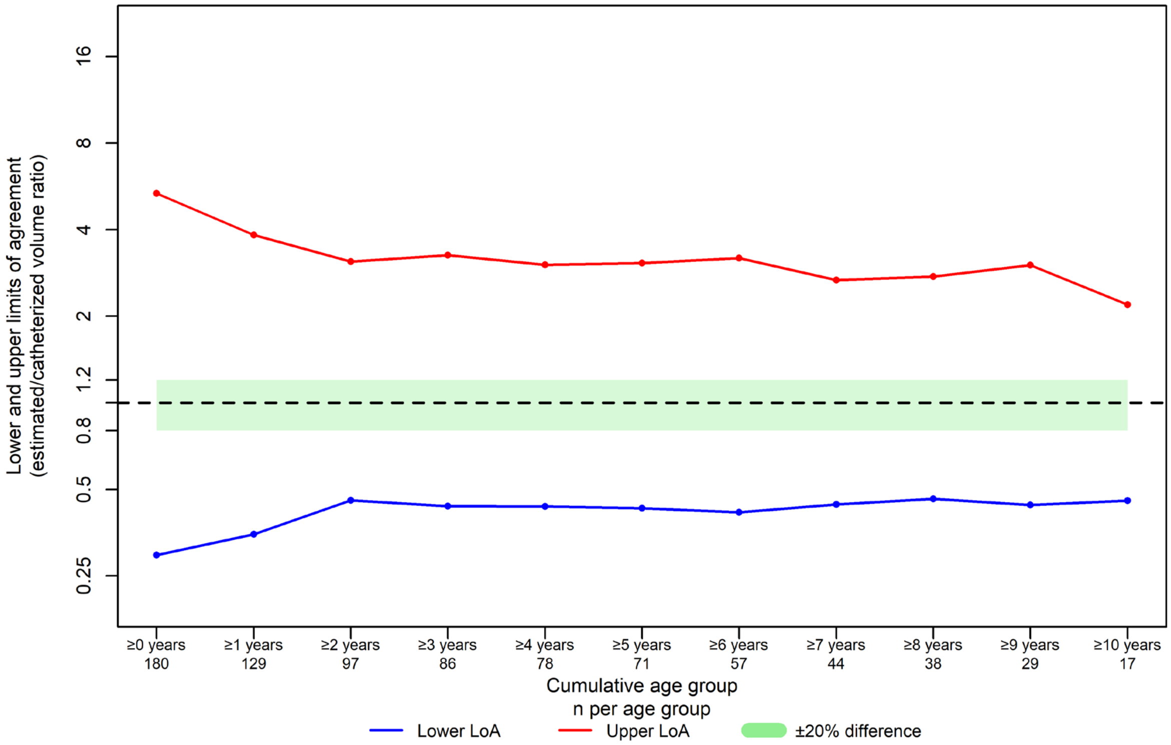


*Cumulative limits of agreement (LoA) progressively narrowed with cumulative age ≥2 years. Across all cumulative age thresholds, the LoA remained outside the predefined ±20% agreement range. Nine measurements with zero values were excluded from the analysis.*

**Supplemental Fig 4. ROC curves for bladder scanner-based detection of urinary retention**

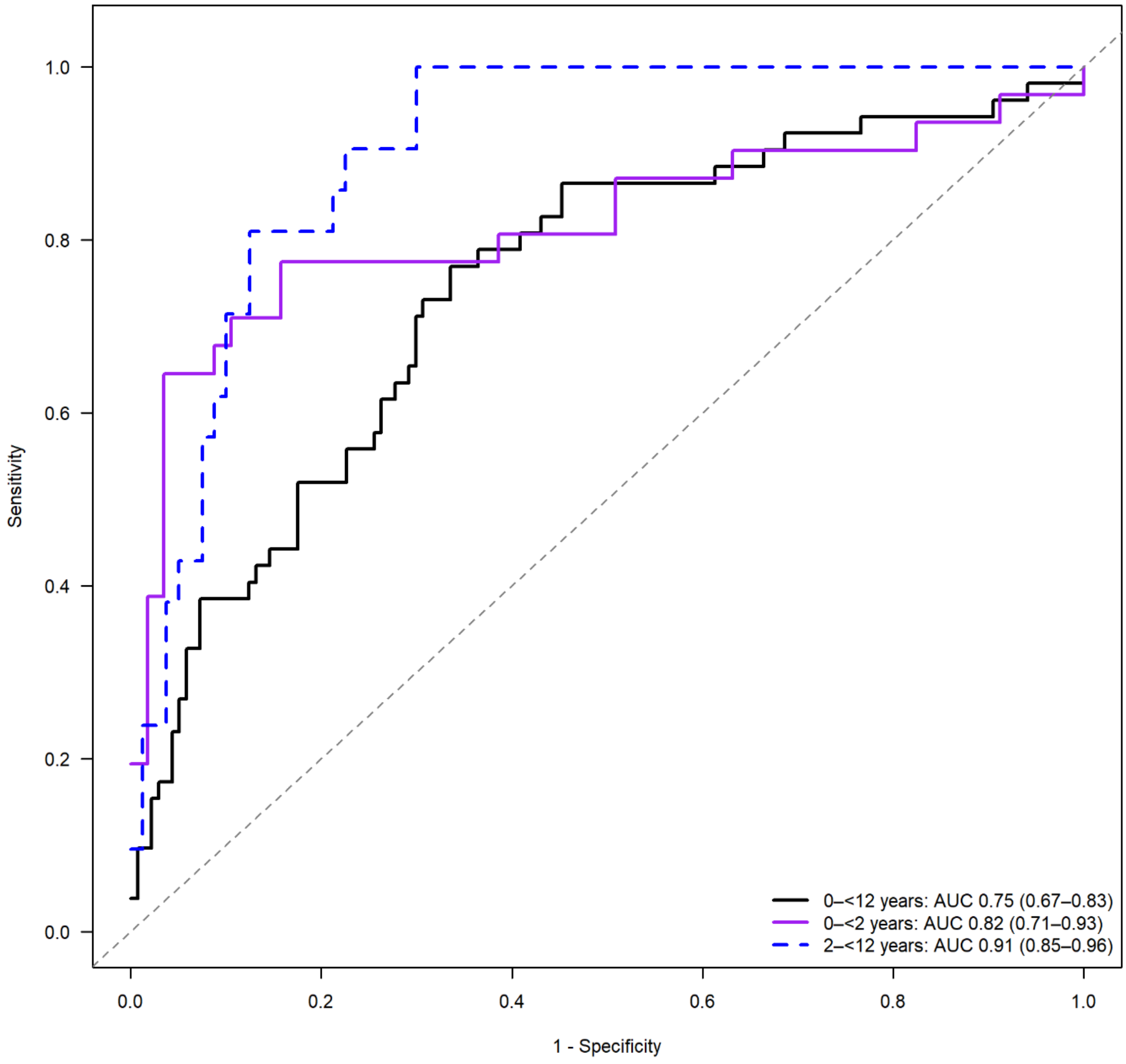


*ROC: Receiver Operator Characteristic curve. Urinary retention was defined as catheterized bladder volume ≥100% expected bladder capacity (EBC): <1 year: [*$weight\ in\ kg \times 7\ mL$*]; ≥ 1 year:* $[(age\ in\ years + 1) \times 30\ mL]$*.*[1,2]

**Supplemental Fig 5. ROC curves for bladder scanner-based detection of elevated post-void residual**

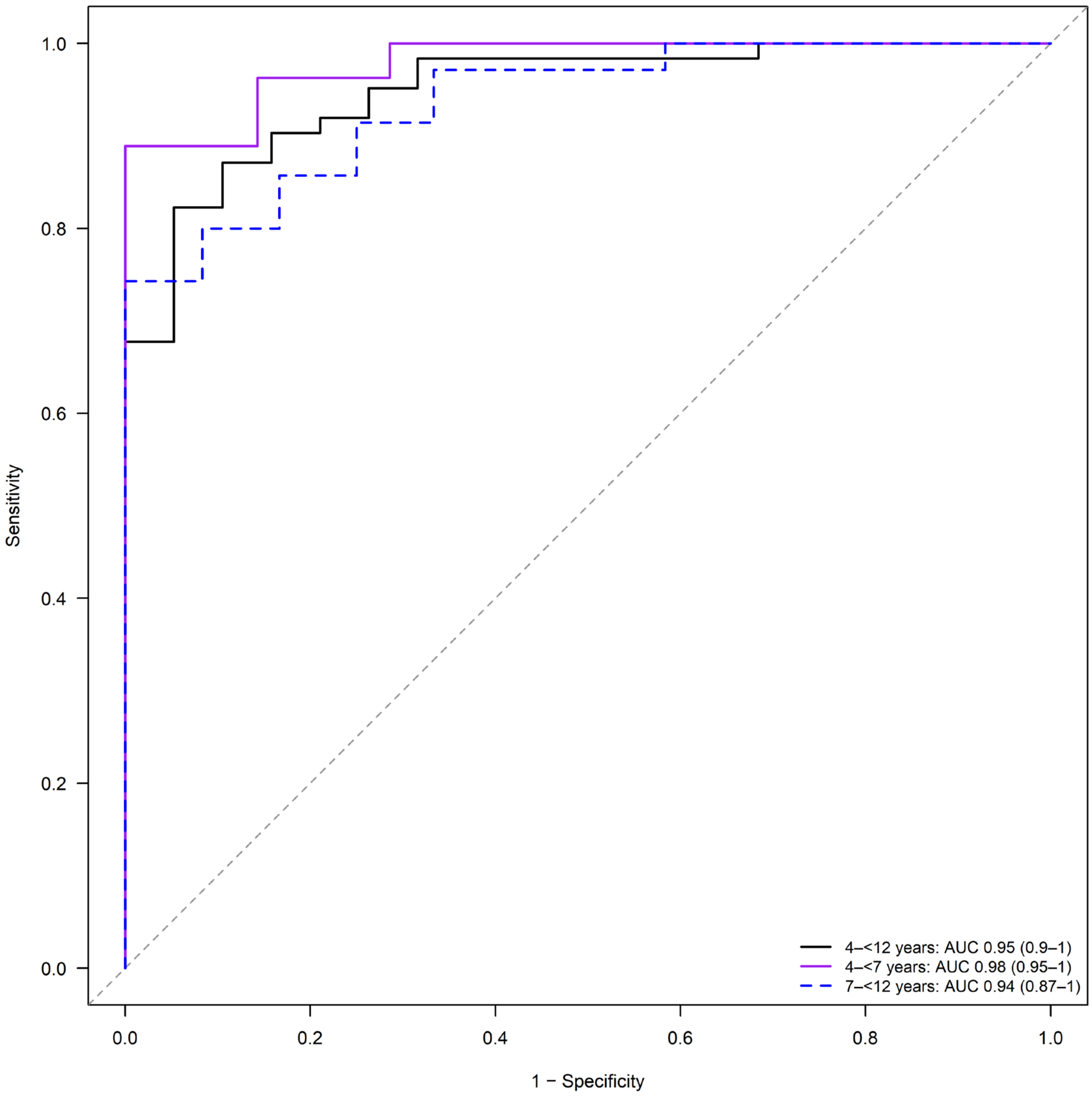


*ROC: Receiver Operator Characteristic curve. Elevated post-void residual (PVR) was defined as >21% expected bladder capacity (EBC) in children aged 4–<7 years (n = 34) and >15% EBC in children aged 7–<12 years (n = 35), irrespective of voiding status.*[2]

### References (supplemental file only)


1. Fairhurst JJ, Rubin CM, Hyde I, Freeman NV, Williams JD. Bladder capacity in infants. *J Pediatr Surg*. Jan 1991;26(1):55–7. doi:10.1016/0022-3468(91)90426-t
2. Austin PF, Bauer SB, Bower W, et al. The standardization of terminology of lower urinary tract function in children and adolescents: Update report from the standardization committee of the International Children's Continence Society. *Neurourol Urodyn*. Apr 2016;35(4):471–81. doi:10.1002/nau.22751
